# Incidence and Risk Factors of Second Primary Cancer after the Initial Primary Human Papillomavirus Related Neoplasms

**DOI:** 10.1101/2020.01.29.19004507

**Authors:** Jiayi Shen, Huaqiang Zhou, Jiaqing Liu, Zhonghan Zhang, Wenfeng Fang, Yunpeng Yang, Shaodong Hong, Yuxiang Ma, Ting Zhou, Yaxiong Zhang, Hongyun Zhao, Yan Huang, Li Zhang

## Abstract

**Background:** Human papillomavirus (HPV)-related cancers are nowadays associated with better survival. However, comprehensive studies in second primary cancer (SPC) after the initial primary HPV-related cancer still remain warranted. Therefore, this study was designed to analyse the incidence and risk factors of SPC after HPV-related cancer.

**Methods:** We identified 86,790 patients diagnosed with initial primary HPV-related cancer between 1973 and 2010 in the National Cancer Institute’s Surveillance, Epidemiology, and End Results (SEER) 9 database. Standardized incidence ratio (SIR) and cumulative incidence were calculated to assess the risk of SPC after HPV-related cancer. Subdistribution hazard regression was also conducted to figure out the risk factors.

**Results:** SIR of SPC after HPV-related cancer was 1.60 (95% confidence interval [CI] 1.55-1.65) for male and 1.25 (95% CI 1.22-1.28) for female. SIR of second primary HPV-related cancer (7.39 [95% CI 6.26-8.68] male and 4.35 [95% CI 4.04-4.67] female) was obviously higher than that of second primary HPV-unrelated cancer (1.54 [95% CI 1.49-1.60] male and 1.16 [95% CI 1.13-1.19] female). 5-year cumulative incidence of SPC was 7.22% (95% CI 6.89%-7.55%) for male and 3.72% (95% CI 3.58%-3.88%) for female. Risk factors of SPC included being married and initial primary cancer (IPC) diagnosed at earlier stage for both gender, and IPC diagnosed at older age as well as surgery performed for female only.

**Conclusion:** Patients diagnosed with HPV-related cancer are more likely to develop another primary cancer, as compared with the age-specific reference population. Patients with the risk factors claimed in this study are suggested to screen for SPC regularly. According to the elevation of SIR of HPV-related SPC, it is suggested that part of the HPV-related SPC cases may be caused by the persistent infection of HPV.

## Introduction

HPV infection is a common viral infection, with its relatively high prevalence (11% in oral cavity, 26.8% for female in genital organs, 14% for female in anus, 45.2% for male in genital organs and 16% for male in anus).^1-5^ Many types of HPV have been classified as Group 1 carcinogen by the International Agency for Research on Cancer (IARC). HPV is a carcinogen in some cancers, with base of tongue, tonsillar, oropharyngeal, Waldeyer’s ring, anal, vulvar, vaginal, cervical and penile cancers included.^6-9^ We grouped cancers listed above as HPV-related cancer and the rest as HPV-unrelated cancer. According to the study by de Martel et al., HPV infection is responsible for 4.5% (630,000 new cases per year) of all cancer cases worldwide. The attributable fraction ranges from < 3% to > 20% in different regions.^10^ Despite the burden of HPV-related cancer, we have seen the improvement in the prognosis of HPV-related cancer, due to the more advanced treatment and earlier detection.^11-16^ Improved survival from the IPC means the elevated risk of SPC.^17^ The investigation into SPC after HPV-related cancer is urgent, based on the high prevalence of HPV-related cancer and the improved survival.

There have been some reports about incidence of SPC after some of the HPV-related cancers. SIR after oral, oropharyngeal, anal and cervical cancer was 2.82, 2.99, 1.41 and 1.56 respectively.^18-20^ However, in the light of the strong carcinogenic effect of HPV, HPV-related cancer should be classified as a whole when considering SPC, to observe whether there will be some special patterns of the occurrence of SPC. Hereby, we aimed to analyse the incidence and risk factors of SPC after initial primary HPV-related cancer.

## Materials and Methods

SEER program has been collecting demographic and clinical characteristics and follow up of cancer patients in America since 1973, with its registries covering approximately 34.6 percent of the U.S. population currently. In this study, we acquired data from SEER 9, a database in SEER*Stat software version 8.3.5 containing information of cancer patients from 9 different registers.

Patient cohort to be observed in this study included those who were diagnosed to have HPV-related cancer as their IPC between 1973 and 2010. IPC was defined as the first record of primary malignant tumor in SEER system. HPV-related cancer included cancer in base of tongue (C01.9), tonsillar cancer (C02.4, C09), oropharyngeal cancer (C10), Waldeyer’s ring cancer (C14.2), anal canal cancer (C21), vulvar cancer (C51), vaginal cancer (C52.9), cervical cancer (C53), and penile cancer (C60).^6-9^ Those diagnosed with HPV-related cancer when aged less than 18 years old were not involved in this study. Records with death certificate or autopsy only were excluded. Finally, 86,790 patients diagnosed with initial primary HPV-related cancer were included.

SPC was defined as the second primary malignant tumor records in SEER system diagnosed between January 1973 and December 2015 and with a latency over or equal to 6 months.^21^ The year of 2010 and 2015 were selected as the last years for the diagnosis of initial primary HPV-related cancer and SPC, respectively, to ensure that all patients included were followed up for at least 5 years.

SIR of SPC is defined as observed number of SPC cases in the study cohort divided by expected number of cases computed using age-specific rates from a reference population, weighted according to the age structure of the study population.^22^ The calculation of SIR was done via the MP-SIR session in SEER software. SIR of SPC after the diagnosed of HPV-related cancer and HPV-unrelated cancer were calculated.

In this study, death is regarded as the competing event with SPC. Cumulative incidence analysis was completed under the consideration of competing risk. To figure out the risk factors of SPC after the initial primary HPV-related cancer, we conducted the multivariable subdistribution hazard regression, regarding death as the competing event with SPC.^23^ Age, race, marital status, TNM stage and surgery were considered to be potential risk factors of SPC. When conducting subdistribution hazard regression, patient cohort merely included those who were diagnosed to have HPV-related cancer as their IPC between 2004 and 2010, for the reason that TNM stage (6th edition) is only available since 2004. Correspondingly, SPC were defined as SPC records in SEER system diagnosed between January 2004 and December 2015.

All the results were presented by gender, except when the IPC was genital cancer, to avoid unexpected bias. Both the cumulative incidence analysis and the subdistribution hazard regression were performed using R version 3.5.0 (Institute for Statistics and Mathematics, Vienna, Austria; www.r-project.org). Statistical significance was set at two-sided P < 0.05.

## Results

### Incidence of SPC

The SIR of SPC after HPV-related cancer was 1.60 (95% CI 1.55-1.65) for male and 1.25 (95% CI 1.22-1.28) for female. (Figure 1) Moreover, SIR of second primary HPV-related cancer (7.39 [95% CI 6.26-8.68] male and 4.35 [95% CI 4.04-4.67] female) was obviously higher than that of second primary HPV-unrelated cancer (1.54 [95% CI 1.49-1.60] male and 1.16 [95% CI 1.13-1.19] female). When SPC site was limited to every single HPV-related site, the consistent elevation of SIR was observed. However, notably, for female patients, SIR of SPC in cervix uteri (1.10 [95% CI 0.89-1.34]) was similar to that of SPC in all sites (1.25 [95% CI 1.22-1.28]). There has been insufficient evidence for the role of HPV in several cancers, which were considered as potentially HPV-related cancer in this study, with esophagus, larynx, nose and nasal sinus, lung, colon and rectum, breast, prostate and urinary bladder included.^24-31^ We also observed an increase in SIR of SPC among some of the potentially HPV-related sites, with esophagus (4.16 [95% CI 3.49-4.93] male and 2.88 [95% CI 2.34-3.51] female), larynx (3.21 [95% CI 2.59-3.94] male and 3.00 [95% CI 2.31-3.82] female), nose and nasal sinus (4.36 [95% CI 2.39-7.32] for male only), lung and bronchus (2.66 [95% CI 2.48-2.84] male and 2.08 [95% CI 1.98-2.18] female), trachea (8.42 [95% CI 1.74-24.61] male and 6.48 [95% CI 2.11-15.13] female) and urinary bladder (1.96 [95% CI 1.74-2.20 for female only) included.

**Figure 1.**
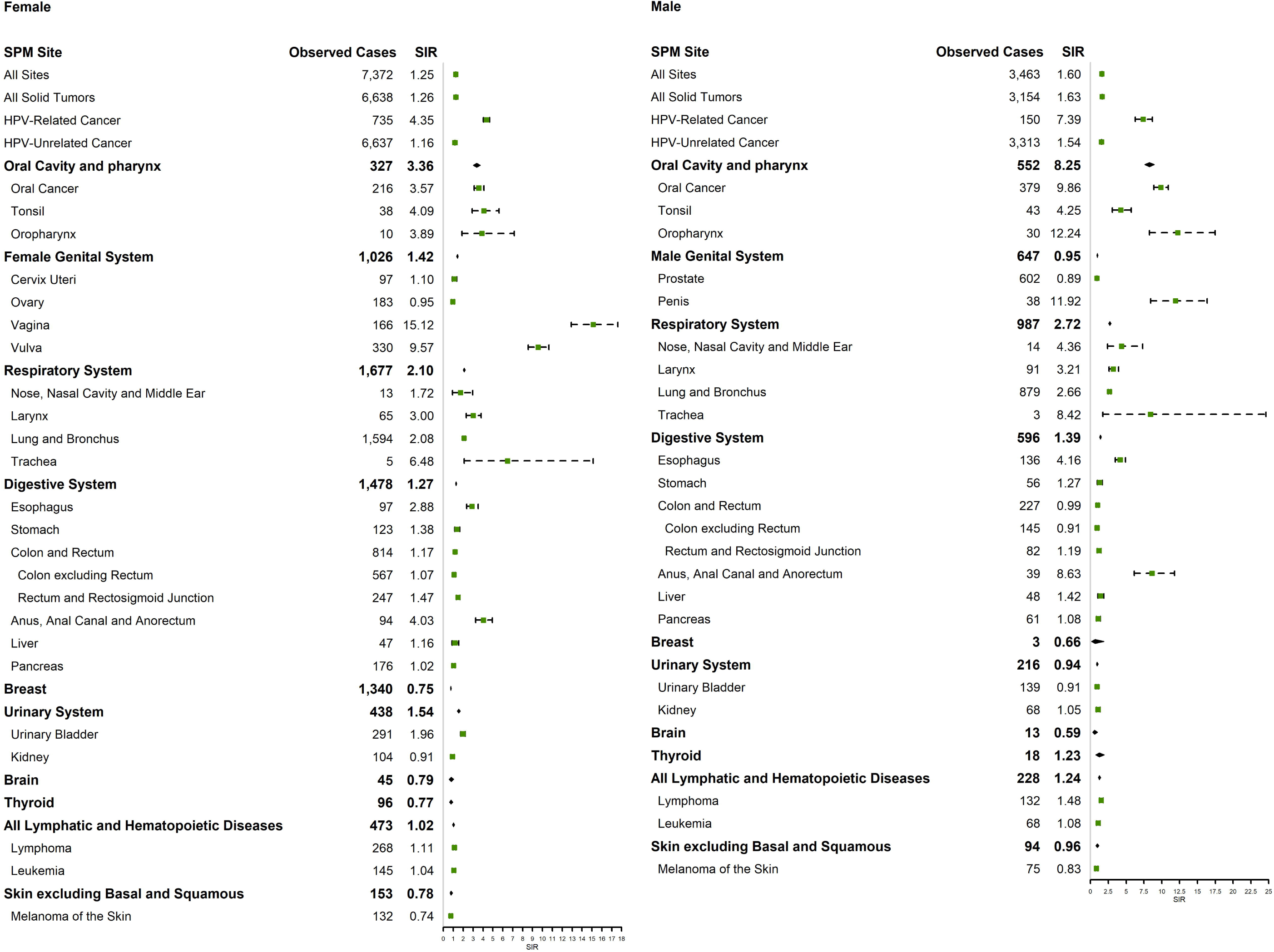
SIRs after the diagnosis of the initial primary HPV-related cancer

HPV-related head and neck cancer contributed more to the SIR of SPC after HPV-related cancer than HPV-related anogenital cancer. (Table 1) In addition, how SIR was affected by SPC sites when IPC was limited to every single HPV-related cancer was also provided. (Table S1-6)

**Table 1.**
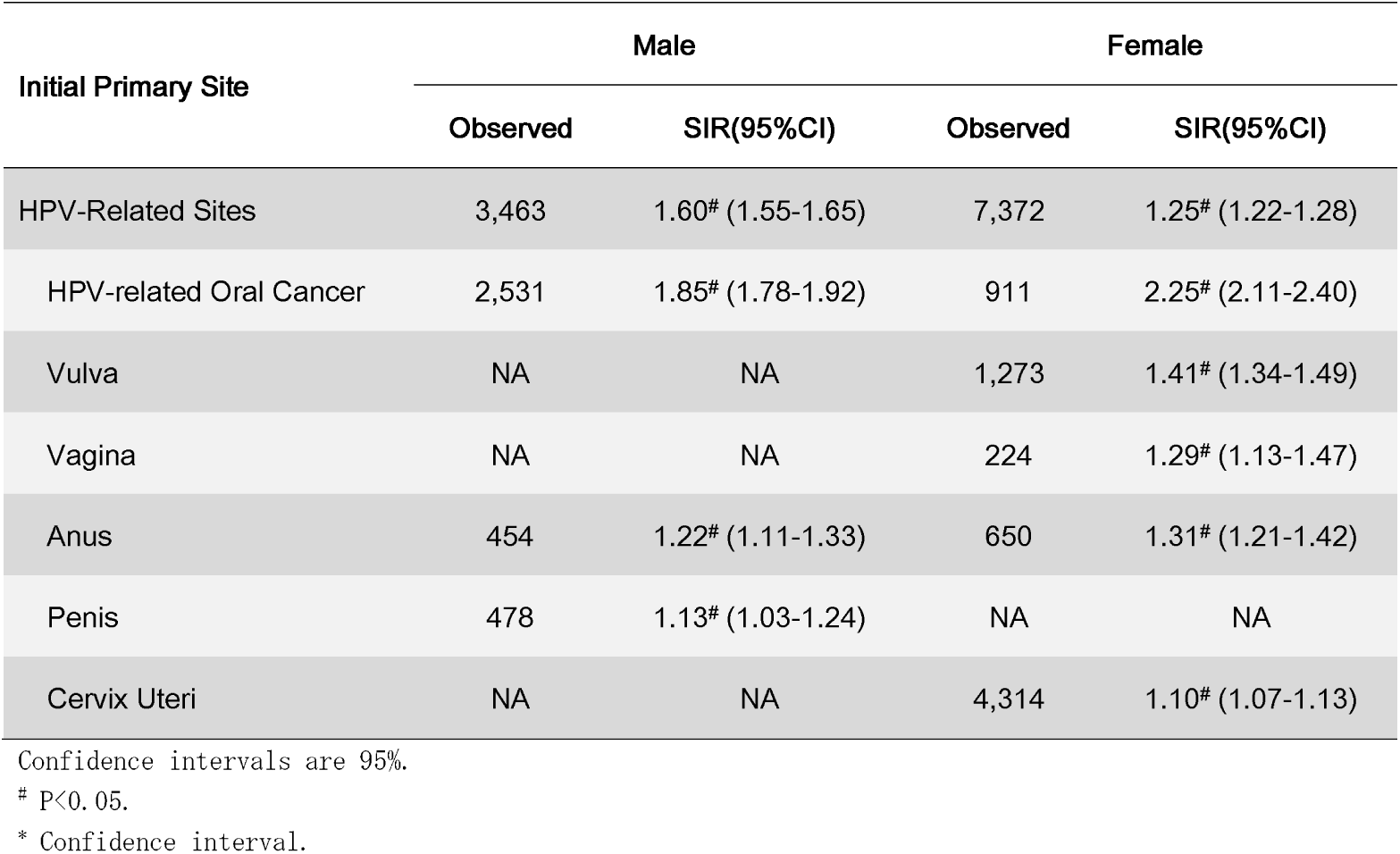
SIRs of all site after the diagnosis of HPV-related cancer (initial primary sites shown)

| Initial Primary Site | Male |  | Female |  |
| --- | --- | --- | --- | --- |
|  | Observed | SIR(95%CI) | Observed | SIR(95%CI) |
| HPV-Related Sites | 3,463 | 1.60<sup>#</sup> (1.55-1.65) | 7,372 | 1.25<sup>#</sup> (1.22-1.28) |
| HPV-related Oral Cancer | 2,531 | 1.85<sup>#</sup> (1.78-1.92) | 911 | 2.25<sup>#</sup> (2.11-2.40) |
| Vulva | NA | NA | 1,273 | 1.41<sup>#</sup> (1.34-1.49) |
| Vagina | NA | NA | 224 | 1.29<sup>#</sup> (1.13-1.47) |
| Anus | 454 | 1.22<sup>#</sup> (1.11-1.33) | 650 | 1.31<sup>#</sup> (1.21-1.42) |
| Penis | 478 | 1.13<sup>#</sup> (1.03-1.24) | NA | NA |
| Cervix Uteri | NA | NA | 4,314 | 1.10<sup>#</sup> (1.07-1.13) |

Confidence intervals are 95%.
<sup>#</sup> P<0.05.
\* Confidence interval.

SIR of SPC after HPV-unrelated cancer (0.88 [95% CI 0.88-0.89] male and 1.12 [95% CI 1.11-1.12] female) was lower when compared with that of SPC after HPV-related cancer (1.60 [95% CI 1.55-1.65] male and 1.25 [95% CI 1.22-1.28] female). (Figure 2)

**Figure 2.**
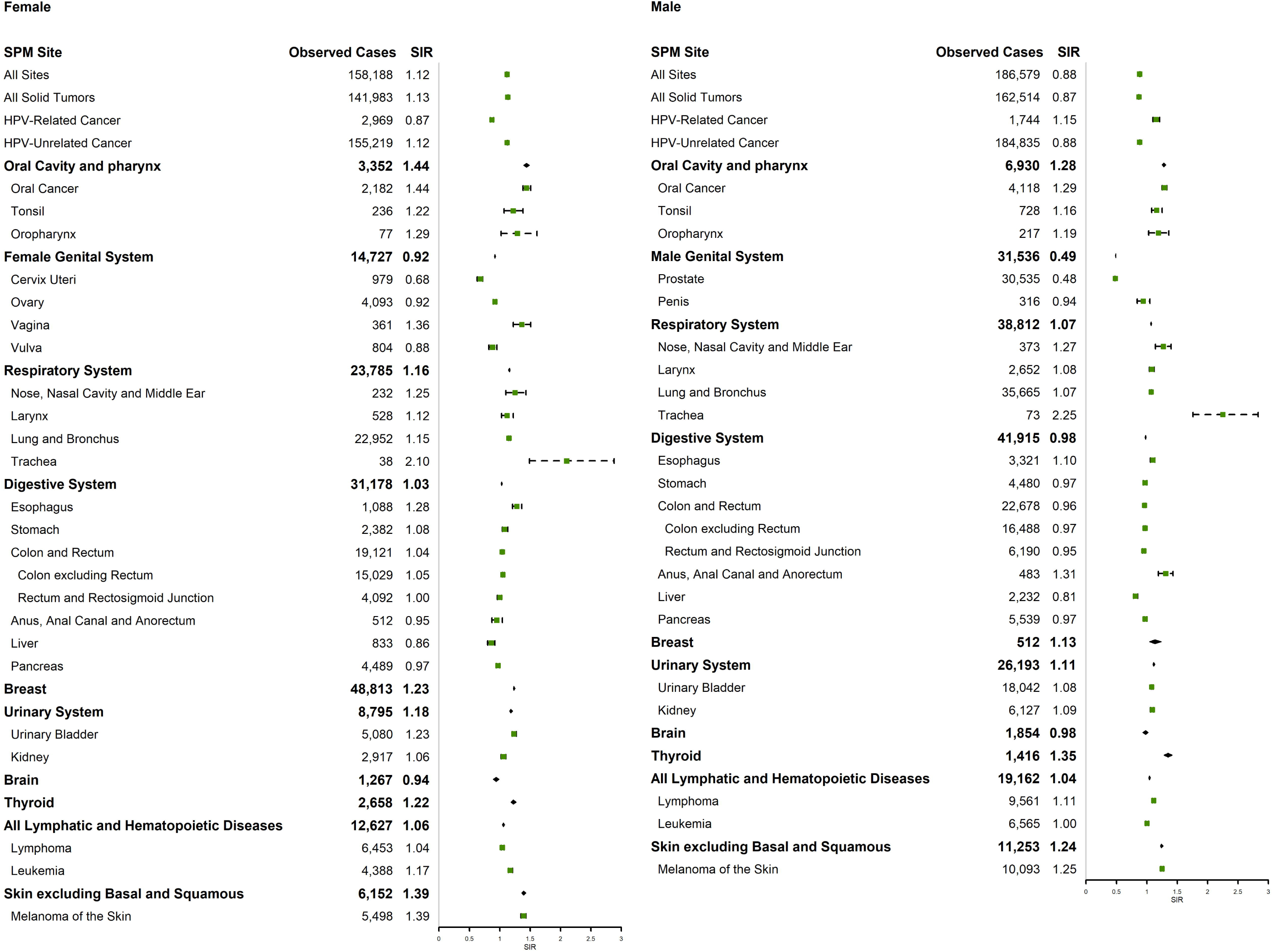
SIRs after the diagnosis of the initial primary HPV-unrelated cancer

The temporal pattern of SIR was apparent. SIR would be raised, if the IPC was diagnosed in more recent years. (Figure 3) The 1-year, 3-year, 5-year and 10-year cumulative incidence of SPC was 1.19% (95% CI 1.06%-1.33%) for male and 0.63% (95% CI 0.57%-0.69%) for female, 4.78% for male (95% CI 4.51%-5.06%) and 2.33% (95% CI 2.22%-2.45%) for female, 7.22% (95% CI 6.89%-7.55%) for male and 3.72% (95% CI 3.58%-3.88%) for female and 11.98% (95% CI 11.56%-12.42%) for male and 6.87% (95% CI 6.67%-7.08%) for female, respectively.

**Figure 3.**
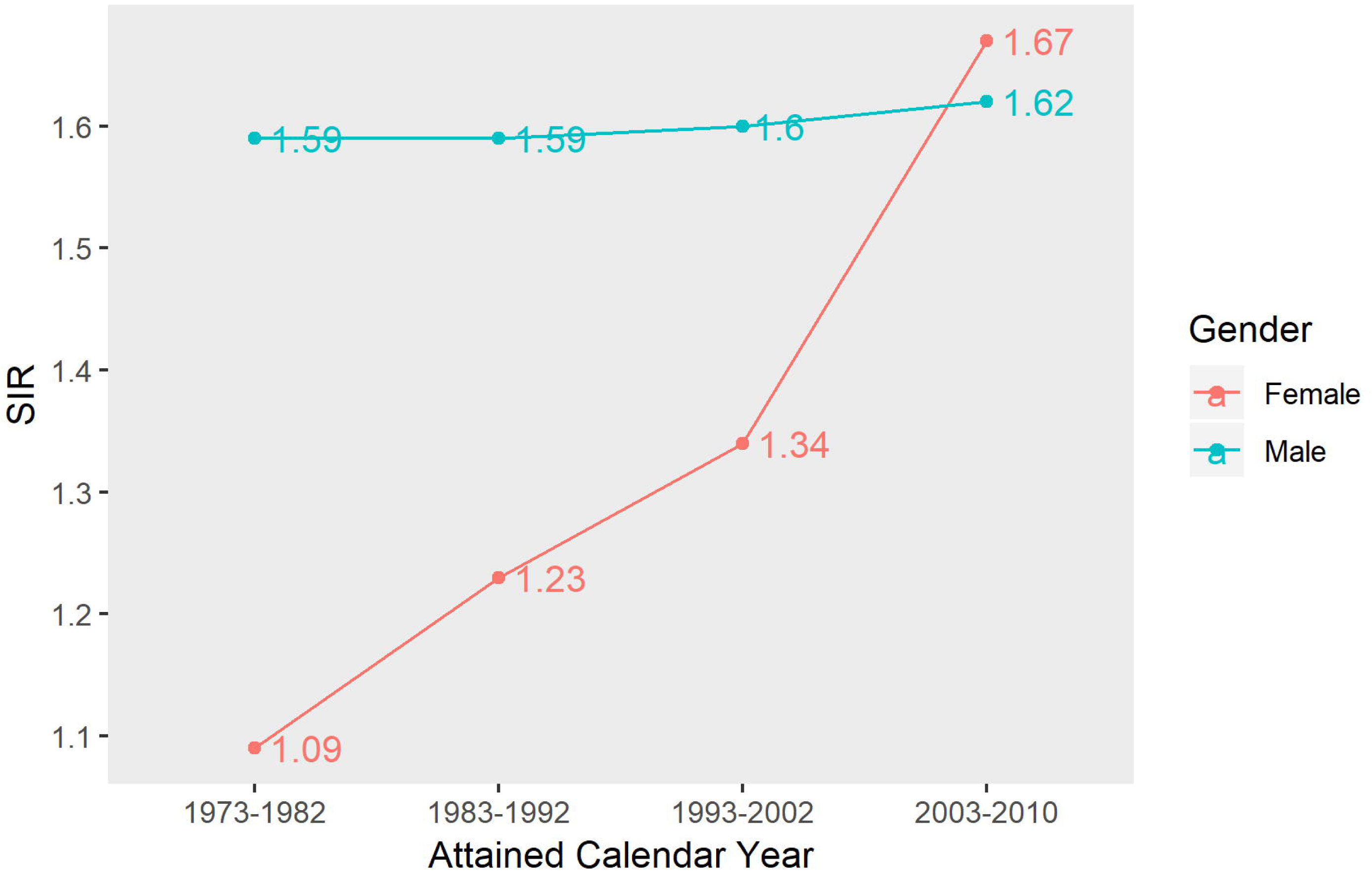
SIRs after the diagnosis of the initial primary HPV-related cancer by attained calendar year

### Risk factors of SPC

Marital status and TNM stage were significantly associated with SPC for both genders. (Table 2) Age and surgery were also significant predictive factors of SPC for female patients.

**Table 2.** Hazard model of probabilities of SPM after initial primary HPV-related cancer.

| Characteristic | SPM of Male |  | SPM of Female |  |
| --- | --- | --- | --- | --- |
|  | sdHR# (95% CI') | p-value | sdHR# (95% CI') | p-value |
| **Age** |  |  |  |  |
| 18-65 years | Reference |  |  |  |
| 65+ years | 1.08 (0.92 - 1.27) | 0.360 | 1.50 (1.28 - 1.76) | <0.001 |
| **Race** |  |  |  |  |
| White | Reference |  |  |  |
| Black | 1.04 (0.82 - 1.32) | 0.730 | 1.08 (0.87 - 1.34) | 0.470 |
| Other/Unknown | 0.74 (0.51 - 1.09) | 0.130 | 0.92 (0.72 - 1.17) | 0.480 |
| **Marital Status** |  |  |  |  |
| Married | Reference |  |  |  |
| Unknown | 0.77 (0.54 - 1.10) | 0.150 | 0.88 (0.66 - 1.17) | 0.390 |
| Unmarried | 0.74 (0.63 - 0.86) | <0.001 | 0.77 (0.66 - 0.90) | <0.001 |
| **TNM stage** |  |  |  |  |
| I | Reference |  |  |  |
| II | 0.74 (0.56 - 1.00) | 0.049 | 1.11 (0.90 - 1.36) | 0.320 |
| III | 0.87 (0.67 - 1.15) | 0.330 | 0.84 (0.67 - 1.05) | 0.120 |
| IV | 0.67 (0.53 - 0.85) | 0.001 | 0.66 (0.50 - 0.86) | 0.002 |
| Unknown | 0.67 (0.50 - 0.91) | 0.009 | 1.10 (0.88 - 1.37) | 0.420 |
| **Surgery** |  |  |  |  |
| Surgery Performed | Reference |  |  |  |
| Surgery not Performed | 0.97 (0.83 - 1.13) | 0.660 | 0.73 (0.62 - 0.86) | <0.001 |
| Unknown | 0.98 (0.46 - 2.11) | 0.960 | 0.14 (0.03 - 0.55) | 0.005 |

According to the result of subdistribution hazard regression, older female patient (with their IPC diagnosed when aged over 65 years old) (sdHR=1.50, [95% CI 1.28 - 1.76], P<0.001) were at more risk for SPC. As compared with married patients, those unmarried got a lower risk of developing SPC, with the sdHR of 0.74 (95% CI 0.63 - 0.86, P<0.001) for male and 0.77 (95% CI 0.66 - 0.90, P<0.001) for female, respectively. In this study, those variables related to poor prognosis, including later TNM stage for both gender and surgery not performed for female, were also protective factors for SPC.

## Discussion

In this study, we estimated the risk and risk factors of SPC after the initial primary HPV-related cancer. The SIR of SPC was 1.60 (95% CI 1.55-1.65) for male and 1.25 (95% CI 1.22-1.28) for female. The SIR, which was larger than 1, indicated that patients diagnosed with HPV-related cancer are more likely to develop another primary cancer, as compared with the age-specific reference population. Patients with the risk factors claimed in the competing risk analysis are suggested to screen for SPC regularly.

The elevation of SIR (>1) can partially be explained by their persistent carcinogenic lifestyle like smoking and alcohol consumption.^32, 33^ In addition, patients once diagnosed with HPV-related cancer may be also those who were genetically susceptible to cancer, thus being more likely to develop another primary cancer than the corresponding reference population.^34^ If those two hypotheses mentioned above were the only two explanations of SPC after HPV-related cancer, we would have seen the similar SIR of different SPC sites, because the carcinogenic effect of both the carcinogenic lifestyle and being genetically susceptible to cancer exists in most of cancers. However, we actually observed the elevation of SIR of SPC in HPV-related sites and this result is also supported by Neumann et al.^35^ Moreover, considering the strong carcinogenic effect of HPV in the HPV-related sites^6-9^, it is suggested that part of the HPV-related SPC cases may be caused by the persistent infection of HPV.

HPV can be transmitted from the primary infected site to the later infected sites. To be specific, HPV virus can circulate to infect other organs, with HPV DNA and mRNA detected in peripheral blood of cervical cancer patients.^36, 37^ Another theory for the transmission of HPV from anus, cervix uteri and other genital organs to oral cavity, oropharynx and tonsil and then causing SPC is that HPV was transmitted in the process of oral sex or by touching the mouth with contaminated hands. Moreover, another primary cancer in the same site as that in IPC may also occur under the effect of HPV, after the elimination of tumor cells of IPC. Next, in those organs infected later, the similar carcinogenic process of HPV with that in IPC occurs. Considering the carcinogenic effect of HPV in both the IPC and its potential carcinogenic effect in second primary HPV-related cancer, HPV vaccine is promoted for the prevention, based on the benefit of HPV vaccination claimed by Nicol et al.^38^

We observed the elevation of SIR in some of the potentially HPV-related SPC sites including esophagus, larynx, nose and nasal sinus (for male only), lung and bronchus, trachea and urinary bladder (for female only). According to the hypothesis that the infection of HPV may be a reason for the occurrence of SPC, we think this can be an indirect evidence for the carcinogenicity of HPV in these potentially HPV-related sites, of which SIR was elevated. For instance, as was promoted by Xiong WM et al., HPV can be transmitted to and infect lung by blood circulation, through the air or by high-risk sexual behavior, through which HPV was transmitted to mouth, throat and then lung.^39^ Then, HPV performs its carcinogenicity, causing second primary lung cancer.^36, 37^ And this is also an indication for the necessity of further exploration of the carcinogenic effect of HPV in esophagus, larynx, nose and nasal sinus, lung and bronchus, trachea and urinary bladder.

The SIR of SPC after HPV-unrelated cancer is lower than 1 for male. But this can not be the evidence of that HPV-unrelated cancer is a protective factor for the development of another primary cancer in male, because there have been many reports of SIR after HPV-unrelated cancers like hepatocellular cancer(10.07), stage Ia non-small-cell lung cancer(2.07), gastric cancer (1.11) and osteosarcoma (1.60), which was significantly larger than 1.^21, 40-42^ However, this can be an indication that there might be survivors from some specific cancers who may be less likely to develop another primary cancer when compared with the corresponding population due to the poor prognosis or so on, which needs further investigation. For example, SIR of SPC after the initial primary gastroenteropancreatic neuroendocrine tumors was reported to be 0.72.^43^

We also saw the increase of SIR by attained calendar year, which means a growing necessity of screening for SPC nowadays. Our results were same to those by Suk et al.^44^ We assumed that the possible reason for the increase may be the improvement of survival from many of the HPV-related cancers in recent years and the arising and popularization of the more effective detection approaches.^11-16, 45, 46^ The improvement of survival reduced the competing effect of death with SPC, while the more effective detection approach made the detection of some cases possible.

According to the result of the subdistribution hazard regression, some prognostic indicators of IPC were also indicators for the occurrence of SPC, with TNM stage (for both gender) and surgery (for female only) included. Further, indicators of better survival of IPC also implied a larger chance of developing SPC, which was supported by the Grundmann et al.^17^

There are also some limitations in our study. First of all, although we believe in the quality of records in SEER system, there may be some incorrect records of SPC in SEER, for the natural difficulty in distinguishing SPC from tumor metastasis and recurrence. Second, information about infection of HPV and lifestyle of patients like smoking and alcohol consumption is not available in SEER database. Because of the lack of information about HPV-positivity status, HPV-relatedness was by proxy in this study.

## Conclusions

This study is of clinical significance and can help promote public health. According to our results, considering the persistent carcinogenesis of HPV and the SIR of SPC increasing by year, patients once diagnosed with HPV-related cancer should screen for SPC regularly, especially when they get a high predictive risk for SPC according to the result of our subdistribution hazard regression.

## Data Availability

The data that support the findings of this study are available in SEER Incidence Database at https://seer.cancer.gov/data/.

## Acknowledgments

The authors acknowledge the efforts of the Surveillance, Epidemiology, and End Results (SEER) Program tumor registries in providing high quality open resources for researchers. The authors would like to thank the editors and the anonymous reviewer for their valuable comments and suggestions to improve the quality of the paper.

## References

1. Candotto V, Lauritano D, Nardone M, et al. HPV infection in the oral cavity: epidemiology, clinical manifestations and relationship with oral cancer. Oral Implantol. 2017;10: 209–220.

2. Dunne EF, Unger ER, Sternberg M, et al. Prevalence of HPV infection among females in the United States. JAMA. 2007;297: 813–819.

3. Hernandez BY, McDuffie K, Zhu X, et al. Anal human papillomavirus infection in women and its relationship with cervical infection. Cancer Epidemiol. Biomarkers Prev. 2005;14: 2550–2556.

4. Han JJ, Beltran TH, Song JW, Klaric J, Choi YS. Prevalence of Genital Human Papillomavirus Infection and Human Papillomavirus Vaccination Rates Among US Adult Men: National Health and Nutrition Examination Survey (NHANES) 2013-2014. JAMA Oncol. 2017;3: 810–816.

5. Nyitray AG, Carvalho da Silva RJ, Baggio ML, et al. Age-specific prevalence of and risk factors for anal human papillomavirus (HPV) among men who have sex with women and men who have sex with men: the HPV in men (HIM) study. J. Infect. Dis. 2011;203: 49–57.

6. Backes DM, Kurman RJ, Pimenta JM, Smith JS. Systematic review of human papillomavirus prevalence in invasive penile cancer. Cancer Causes Control. 2009;20: 449–457.

7. Vuyst HD, De Vuyst H, Clifford GM, Nascimento MC, Madeleine MM, Franceschi S. Prevalence and type distribution of human papillomavirus in carcinoma and intraepithelial neoplasia of the vulva, vagina and anus: A meta-analysis. International Journal of Cancer. 2009;124: 1626–1636.

8. Walboomers JM, Jacobs MV, Manos MM, et al. Human papillomavirus is a necessary cause of invasive cervical cancer worldwide. J. Pathol. 1999;189: 12–19.

9. Chaturvedi AK, Engels EA, Anderson WF, Gillison ML. Incidence trends for human papillomavirus-related and -unrelated oral squamous cell carcinomas in the United States. J. Clin. Oncol. 2008;26: 612–619.

10. Martel Cd, de Martel C, Plummer M, Vignat J, Franceschi S. Worldwide burden of cancer attributable to HPV by site, country and HPV type. International Journal of Cancer. 2017;141: 664–670.

11. Akhtar-Danesh N, Elit L, Lytwyn A. Temporal Trends in the Relative Survival Among Women With Cervical Cancer in Canada: A Population-Based Study. International Journal of Gynecologic Cancer. 2012;22: 1208–1213.

12. Arya M, Li R, Pegler K, et al. Long-term trends in incidence, survival and mortality of primary penile cancer in England. Cancer Causes Control. 2013;24: 2169–2176.

13. Chen AY, Zhu J, Fedewa S. Temporal trends in oropharyngeal cancer treatment and survival: 1998–2009. The Laryngoscope. 2014;124: 131–138.

14. Johnson LG, Madeleine MM, Newcomer LM, Schwartz SM, Daling JR. Anal cancer incidence and survival: the surveillance, epidemiology, and end results experience, 1973-2000. Cancer. 2004;101:281–288.

15. Lai J, Elleray R, Nordin A, et al. Vulval cancer incidence, mortality and survival in England: age-related trends. BJOG. 2014;121: 728-738; discussion 739.

16. Pulte D, Brenner H. Changes in Survival in Head and Neck Cancers in the Late 20th and Early 21st Century: A Period Analysis. The Oncologist. 2010;15: 994–1001.

17. Grundmann RT, Meyer F. [Second primary malignancy among cancer survivors - epidemiology, prognosis and clinical relevance]. Zentralbl. Chir. 2012;137: 565–574.

18. Morris LGT, Sikora AG, Patel SG, Hayes RB, Ganly I. Second primary cancers after an index head and neck cancer: subsite-specific trends in the era of human papillomavirus-associated oropharyngeal cancer. J. Clin. Oncol. 2011;29: 739–746.

19. Shah BK, Budhathoki N. Second primary malignancies in anal carcinoma. Journal of Clinical Oncology. 2015;33: 534–534.

20. Teng C-J, Huon L-K, Hu Y-W, et al. Secondary Primary Malignancy Risk in Patients With Cervical Cancer in Taiwan. Medicine. 2015;94: e1803.

21. Shah BK, Kandel P, Khanal A. Second Primary Malignancies in Hepatocellular Cancer - A US Population-based Study. Anticancer Res. 2016;36: 3511–3514.

22. Schoenberg BS, Myers MH. Statistical methods for studying multiple primary malignant neoplasms. Cancer. 1977;40: 1892–1898.

23. Fine JP, Gray RJ. A Proportional Hazards Model for the Subdistribution of a Competing Risk. J. Am. Stat. Assoc. 1999;94: 496–509.

24. Kreimer AR, Clifford GM, Boyle P, Franceschi S. Human papillomavirus types in head and neck squamous cell carcinomas worldwide: a systematic review. Cancer Epidemiol. Biomarkers Prev. 2005;14: 467–475.

25. Lu XM, Monnier-Benoit S, Mo LZ, et al. Human papillomavirus in esophageal squamous cell carcinoma of the high-risk Kazakh ethnic group in Xinjiang, China. European Journal of Surgical Oncology (EJSO). 2008;34: 765–770.

26. El-Mofty SK, Lu DW. Prevalence of high-risk human papillomavirus DNA in nonkeratinizing (cylindrical cell) carcinoma of the sinonasal tract: a distinct clinicopathologic and molecular disease entity. Am. J. Surg. Pathol. 2005;29: 1367–1372.

27. Giuliani L, Favalli C, Syrjanen K, Ciotti M. Human papillomavirus infections in lung cancer. Detection of E6 and E7 transcripts and review of the literature. Anticancer Res. 2007;27: 2697–2704.

28. Bodaghi S. Colorectal Papillomavirus Infection in Patients with Colorectal Cancer. Clinical Cancer Research. 2005;11: 2862–2867.

29. Akil N, Yasmeen A, Kassab A, Ghabreau L, Darnel AD, Al Moustafa AE. High-risk human papillomavirus infections in breast cancer in Syrian women and their association with Id-1 expression: a tissue microarray study. Br. J. Cancer. 2008;99: 404–407.

30. Leiros GJ, Galliano SR, Sember ME, Kahn T, Schwarz E, Eiguchi K. Detection of human papillomavirus DNA and p53 codon 72 polymorphism in prostate carcinomas of patients from Argentina. BMC Urology. 2005;5.

31. Barghi MR, Hajimohammadmehdiarbab A, Moghaddam SH, Kazemi B. Correlation between human papillomavirus infection and bladder transitional cell carcinoma. BMC Infectious Diseases. 2005;5.

32. Vineis P, Alavanja M, Buffler P, et al. Tobacco and Cancer: Recent Epidemiological Evidence. JNCI Journal of the National Cancer Institute. 2004;96: 99–106.

33. Boffetta P, Hashibe M. Alcohol and cancer. Lancet Oncol. 2006;7: 149–156.

34. Dong LM, Potter JD, White E, Ulrich CM, Cardon LR, Peters U. Genetic susceptibility to cancer: the role of polymorphisms in candidate genes. JAMA. 2008;299: 2423–2436.

35. Neumann F, Jégu J, Mougin C, et al. Risk of second primary cancer after a first potentially-human papillomavirus-related cancer: A population-based study. Preventive Medicine. 2016;90: 52–58.

36. Tseng C-J, Pao CC, Lin J-D, Soong Y-K, Hong J-H, Hsueh S. Detection of Human Papillomavirus Types 16 and 18 mRNA in Peripheral Blood of Advanced Cervical Cancer Patients and Its Association With Prognosis. Journal of Clinical Oncology. 1999;17: 1391–1391.

37. Liu VW, Tsang P, Yip A, Ng TY, Wong LC, Ngan HY. Low incidence of HPV DNA in sera of pretreatment cervical cancer patients. Gynecol. Oncol. 2001;82: 269–272.

38. Nicol AF, de Andrade CV, Russomano FB, et al. HPV vaccines: their pathology-based discovery, benefits, and adverse effects. Ann. Diagn. Pathol. 2015;19: 418–422.

39. Xiong W-M, Xu Q-P, Li X, Xiao R-D, Cai L, He F. The association between human papillomavirus infection and lung cancer: a system review and meta-analysis. Oncotarget. 2017;8: 96419–96432.

40. Khanal A, Lashari BH, Kruthiventi S, et al. The risk of second primary malignancy in patients with stage Ia non-small cell lung cancer: a U.S. population-based study. Acta Oncologica. 2018;57: 239–243.

41. Shah BK, Khanal A, Hewett Y. Second Primary Malignancies in Adults with Gastric Cancer - A US Population-Based Study. Front. Oncol. 2016;6: 82.

42. Lee JS, DuBois SG, John Boscardin W, Wustrack RL, Goldsby RE. Secondary malignant neoplasms among children, adolescents, and young adults with osteosarcoma. Cancer. 2014;120: 3987–3993.

43. Kamp K, Damhuis RAM, Feelders RA, de Herder WW. Occurrence of second primary malignancies in patients with neuroendocrine tumors of the digestive tract and pancreas. Endocrine-Related Cancer. 2012;19: 95–99.

44. Suk R, Mahale P, Sonawane K, et al. Trends in Risks for Second Primary Cancers Associated With Index Human Papillomavirus–Associated Cancers. JAMA Network Open. 2018;1: e181999.

45. Hillemanns P, Soergel P, Hertel H, Jentschke M. Epidemiology and Early Detection of Cervical Cancer. Oncol Res Treat. 2016;39: 501–506.

46. Leeds IL. Anal cancer and intraepithelial neoplasia screening: A review. World Journal of Gastrointestinal Surgery. 2016;8: 41.

